# CovidCounties - an interactive, real-time tracker of the COVID-19 pandemic at the level of US counties

**DOI:** 10.1101/2020.04.28.20083279

**Authors:** Douglas Arneson, Matthew Elliott, Arman Mosenia, Boris Oskotsky, Rohit Vashisht, Travis Zack, Paul Bleicher, Atul J. Butte, Vivek A. Rudrapatna

## Abstract

Management of the COVID-19 pandemic has proven to be a significant challenge to policy makers. This is in large part due to uneven reporting and the absence of open-access visualization tools to present local trends and infer healthcare needs. Here we report the development of CovidCounties.org, an interactive web application that depicts daily disease trends at the level of US counties using time series plots and maps. This application is accompanied by a manually curated dataset that catalogs all major public policy actions made at the state-level, as well as technical validation of the primary data. Finally, the underlying code for the site is also provided as open source, enabling others to validate and learn from this work.

## Introduction

The disease known as COVID-19 was first reported in December of 2019 in Wuhan, China^1^. Three months later it was declared a pandemic by the WHO, and since then its death toll has reached over 150,000 while infecting over 2 million people across 210 countries worldwide^2^. Additionally, the pandemic has disrupted the daily lives of billions and has incurred significant socioeconomic costs at the global level.

In the US, the very assessment of the disease’s impact has been challenged by limitations in accurate data capture and analysis. Variable testing, uneven reporting, barriers to data sharing, and a lack of easy-to-use analytic tools have all contributed to a lack of clarity in establishing and trending the state of the pandemic. As a consequence, policy makers at all levels have been forced to make decisions of great socioeconomic consequence in the face of significant uncertainty.

To improve the accessibility of basic COVID-19-related information in the US, especially by the general public and policymakers without a data science background, we report the creation of a new interactive visualization tool that depicts daily disease trends at the level of individual US counties. This web application features the novel reuse of several publicly available sources of data while also introducing a new, manually curated dataset accompanying this manuscript. This site features several unique views, including local doubling times and estimated ICU bed requirements by county. Additionally, we report the technical validation of the primary data (counts per county per day) against other official- and commonly used sources of data.

## Methods

### Data sources

Data on state-wide and county-level counts were obtained from The New York Times^3^ via their *github* repository (https://github.com/nytimes/covid-19-data). County-wise population data were obtained from the US Census^4^ using the *R* package *tidycensus*^5^. Data on ICU bed availability per county was obtained from Kaiser Health News^6^.

As per The New York Times, cases and deaths reported from New York, Kings, Queens, Bronx and Richmond counties were assigned to New York City. Similarly, Cass, Clay, Jackson and Platte counties in Missouri were assigned to Kansas City. When a patient’s county of residence was unknown or pending many state departments reported these cases as coming from “unknown” counties. Cases reported from unknown counties were only included at the state level.

Data related to state-wide implementation of social-distancing policies were manually curated by web search and independently reviewed by a second author; disagreements were rare and resolved by discussion. Government websites were prioritized as sources of truth where feasible; otherwise, news reports covering state-wide proclamations were used. All citations are captured in the open data file accompanying this manuscript. [https://datadryad.org/stash/share/whGecW9DWYmoAVMDdAHNF0z712Vbxrj9YwI5QKRAWUs]. These data were up to date and confirmed as of the date of data deposit: April 19, 2020.

Ground truth data used for validation were manually curated from the websites of multiple state departments of public health as well as Corona Data Scraper [https://coronadatascraper.com/], a commonly used resource for aggregating county-level tracking of COVID-19 over time. Citations of the validation data are included in the data file accompanying this manuscript. [https://datadryad.org/stash/share/whGecW9DWYmoAVMDdAHNF0z712Vbxrj9YwI5QKRAWUs]

Descriptive statistics on all datasets except that of the US Census and validation data are reported in Table 1.

**Table 1:**
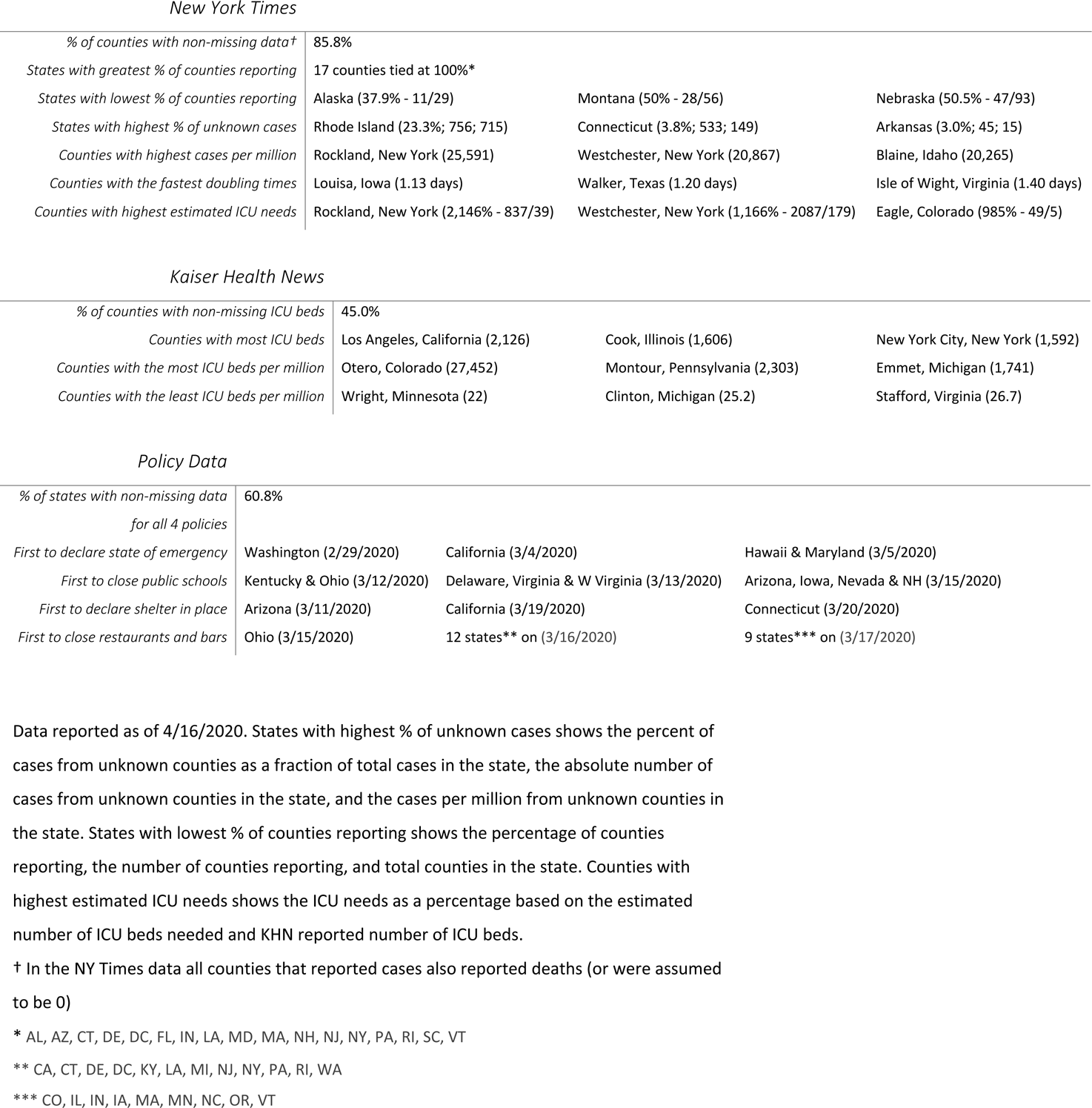
Descriptive statistics on the included data sets.

*New York Times*
|  |  |  |  |
| --- | --- | --- | --- |
| *% of counties with non-missing data†* | 85.8% |  |  |
| *States with greatest % of counties reporting* | 17 counties tied at 100%* |  |  |
| *States with lowest % of counties reporting* | Alaska (37.9% - 11/29) | Montana (50% - 28/56) | Nebraska (50.5% - 47/93) |
| *States with highest % of unknown cases* | Rhode Island (23.3%; 756; 715) | Connecticut (3.8%; 533; 149) | Arkansas (3.0%; 45; 15) |
| *Counties with highest cases per million* | Rockland, New York (25,591) | Westchester, New York (20,867) | Blaine, Idaho (20,265) |
| *Counties with the fastest doubling times* | Louisa, Iowa (1.13 days) | Walker, Texas (1.20 days) | Isle of Wight, Virginia (1.40 days) |
| *Counties with highest estimated ICU needs* | Rockland, New York (2,146% - 837/39) | Westchester, New York (1,166% - 2087/179) | Eagle, Colorado (985% - 49/5) |
| *Kaiser Health News* |  |  |  |
| *% of counties with non-missing ICU beds* | 45.0% |  |  |
| *Counties with most ICU beds* | Los Angeles, California (2,126) | Cook, Illinois (1,606) | New York City, New York (1,592) |
| *Counties with the most ICU beds per million* | Otero, Colorado (27,452) | Montour, Pennsylvania (2,303) | Emmet, Michigan (1,741) |
| *Counties with the least ICU beds per million* | Wright, Minnesota (22) | Clinton, Michigan (25.2) | Stafford, Virginia (26.7) |
| *Policy Data* |  |  |  |
| *% of states with non-missing data for all 4 policies* | 60.8% |  |  |
| *First to declare state of emergency* | Washington (2/29/2020) | California (3/4/2020) | Hawaii & Maryland (3/5/2020) |
| *First to close public schools* | Kentucky & Ohio (3/12/2020) | Delaware, Virginia & W Virginia (3/13/2020) | Arizona, Iowa, Nevada & NH (3/15/2020) |
| *First to declare shelter in place* | Arizona (3/11/2020) | California (3/19/2020) | Connecticut (3/20/2020) |
| *First to close restaurants and bars* | Ohio (3/15/2020) | 12 states** on (3/16/2020) | 9 states*** on (3/17/2020) |

Data reported as of 4/16/2020. States with highest % of unknown cases shows the percent of cases from unknown counties as a fraction of total cases in the state, the absolute number of cases from unknown counties in the state, and the cases per million from unknown counties in the state. States with lowest % of counties reporting shows the percentage of counties reporting, the number of counties reporting, and total counties in the state. Counties with highest estimated ICU needs shows the ICU needs as a percentage based on the estimated number of ICU beds needed and KHN reported number of ICU beds.
† In the NY Times data all counties that reported cases also reported deaths (or were assumed to be 0)
\* AL, AZ, CT, DE, DC, FL, IN, LA, MD, MA, NH, NJ, NY, PA, RI, SC, VT
\*\* CA, CT, DE, DC, KY, LA, MI, NJ, NY, PA, RI, WA
\*\*\* CO, IL, IN, IA, MA, MN, NC, OR, VT

### Doubling Time

Doubling time was calculated for each state and county by taking the reciprocal of difference between the log (base 2) case counts corresponding to adjacent days, then applying the *R* function *loess* for smoothing. The input of this model required a minimum of 8 days of data where the minimum number of cases was greater than 10. Regularization was performed by replacing extreme doubling times (>500 days) with the average of the surrounding values.

### ICU Bed Occupancy Model

We incorporated parameters related to rates of hospitalization and ICU admission from work previously published by Ferguson et al.^7^. Although simpler than other models, it fit publicly available county-level ICU bed data in California well and was easier to understand for the user than more complicated models proposed ^8–11^. This model assumed a 4.4% rate of hospitalization among all new cases, a 30% rate of intensive care unit admission among hospitalized patients, and a 9-day average length of stay (time until discharge or death).

### Web Application Development and Deployment

See Figure 1 for an overall schematic of the web application. The source code was written in *R* (4.1.0)^12^ using the *shiny*^13^*, shinyjs*^14^*, tidyverse*^15^ and *plotly*^16^ packages. Software version control was achieved using Docker. The entire software code for the site is publicly available on *github* (https://github.com/vivical/ButteLabCOVID) and *dockerhub* (https://hub.docker.com/r/pupster90/covid_tracker). The web hosting was organized as a unified data share between all instances running *R shiny* code and controlled by a load balancer using an auto-scaling mechanism. The web environment is hosted by Amazon Web Services and is located at covidcounties.org.

**Figure 1:**
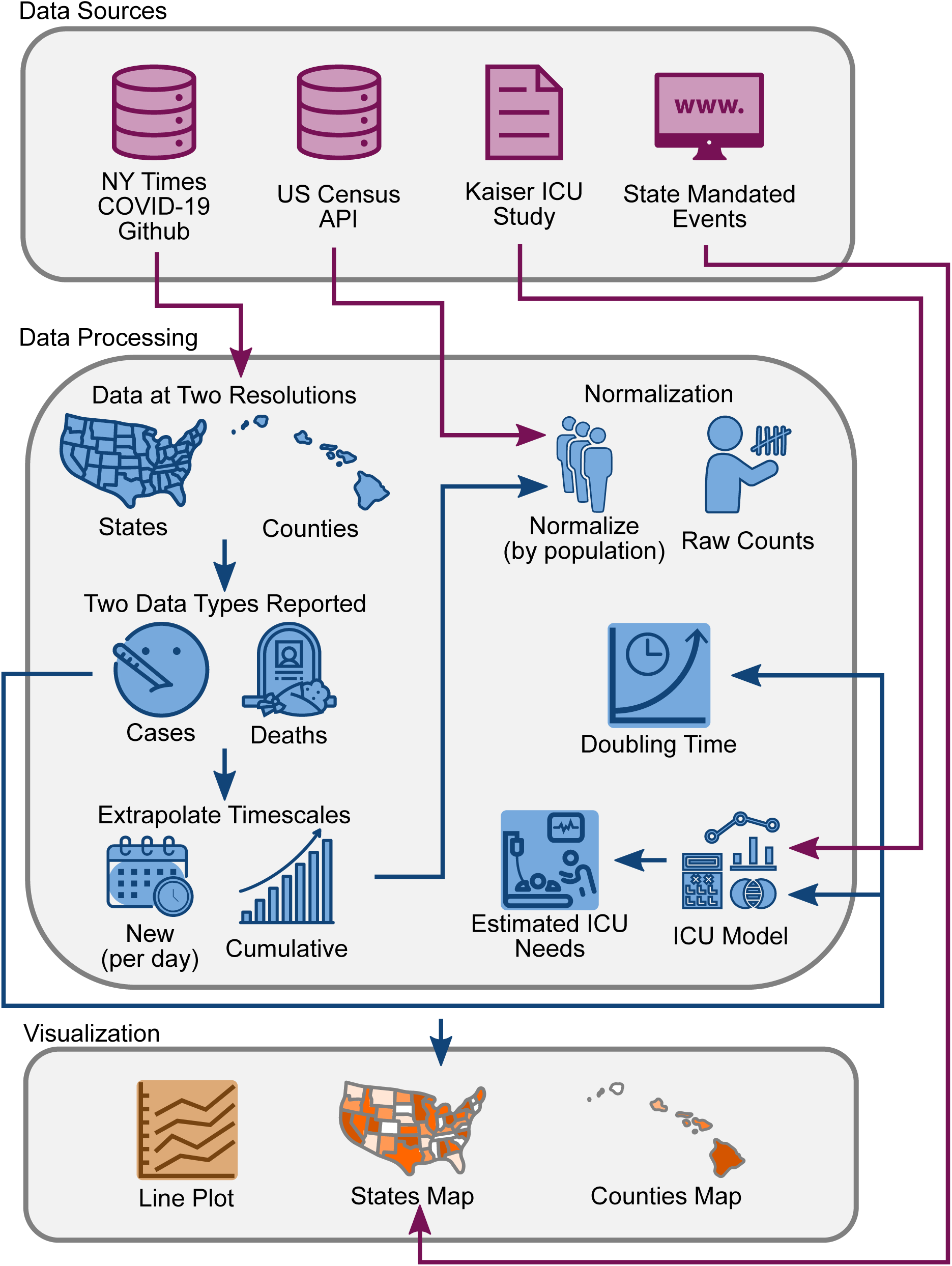
Database schematic. Source data was obtained from The New York Times, US Census Bureau, Kaiser Health News, and from a manual curation of state governmental websites and news outlets as described in *Methods*. Data was processed to reflect case and death counts at the level of states and counties. Functions were written to perform x- and y-axis rescaling, normalization by population, doubling time estimation, and ICU bed utilization. Results were depicted using interactive line plots and maps.

## Results

CovidCounties derives a majority of its data from The New York Times Coronavirus github page [https://github.com/nytimes/covid-19-data] which is updated daily with cases and deaths reported in each state and county from the previous day. This time series dataset was derived from a variety of governmental sources. However, to our knowledge this data has never been formally validated against other reputed sources of COVID-19 reporting including state and local departments of public health.

First, we demonstrate the high concordance of cumulative cases and deaths calculated and displayed in CovidCounties at the county level by directly comparing these to numbers reported by the Departments of Public Health in California and Connecticut (Figure 2A, 2B). These two states were chosen because they both publicly report the daily counts of cases requiring hospitalization or intensive care at the county level. R^2^ rates corresponding to the concordance between predicted and actual counts ranged from 0.86 to 1. To our knowledge, California is only state in the US to report county-wide ICU bed utilization rates. We found a high degree of concordance (R^2^ = 0.87) with minimal model bias (Figure 2A), indicating a fairly high degree of explained variation despite a relatively simplistic model.

**Figure 2:**
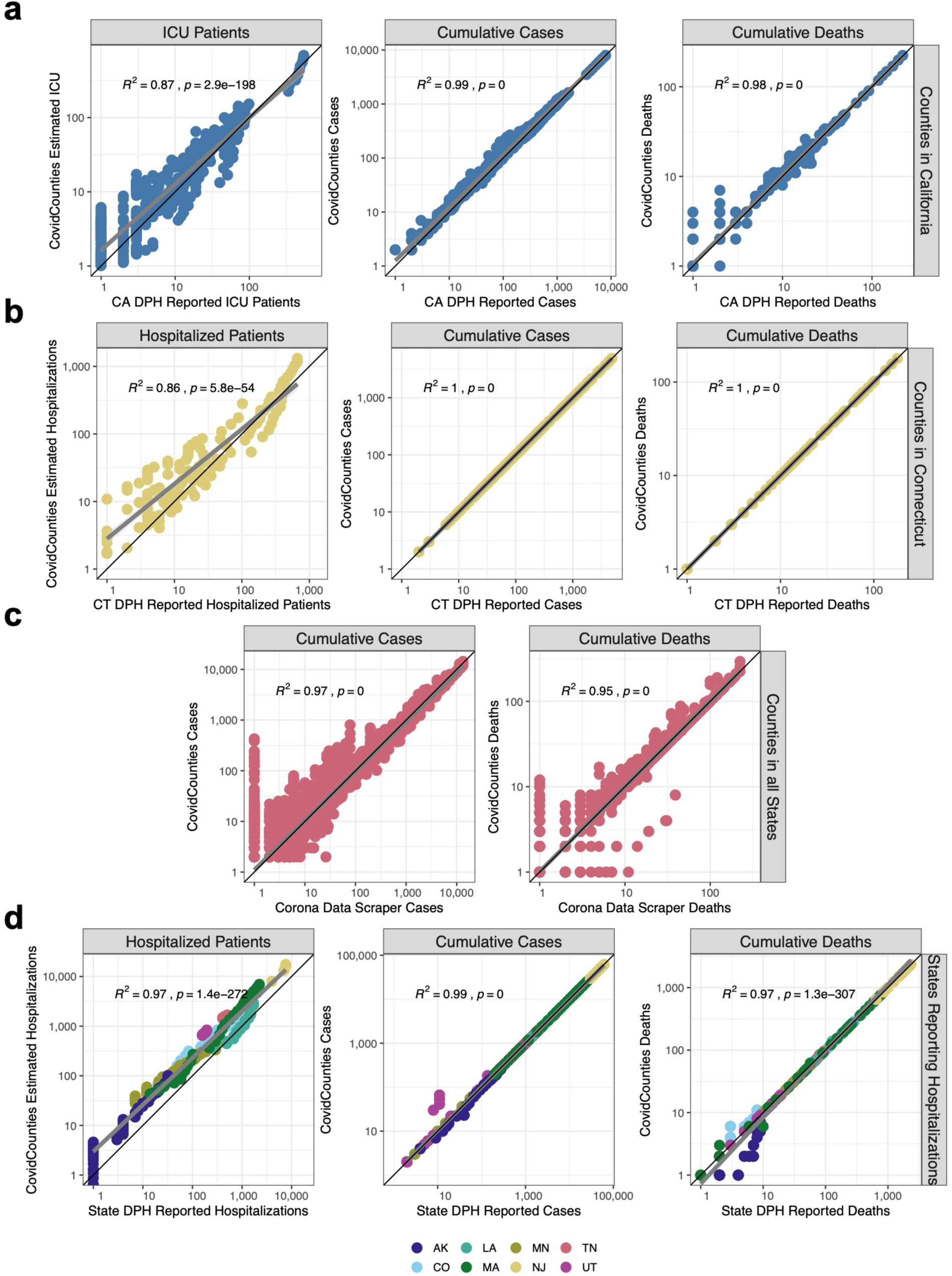
Technical validation of the datasets. A. Comparison of estimated ICU bed occupancy, cumulative cases, and cumulative deaths reported by CovidCounties against corresponding data reported by the California Department of Public Health. Each point corresponds to a measurement from a given California county on a particular date where both datasets report counts. Data is from 4/1/2020 - 4/9/2020. B. Comparison of the estimated hospital bed occupancy, cumulative cases, and cumulative deaths reported by CovidCounties against corresponding data reported by the Connecticut Department of Public Health. Each point corresponds to a measurement from a given Connecticut county on a particular date where both datasets report counts. Data is from 3/24/2020 - 4/9/2020. C. Comparison of the estimated hospital bed occupancy, cumulative cases, and cumulative deaths reported by CovidCounties against corresponding data reported by the website Corona Data Scraper as of 4/10/2020 (includes data up to 4/9/2020). Each point corresponds to a measurement from any US county in the dataset at a particular time where both datasets report counts. D. Comparison of the estimated hospital bed occupancy, cumulative cases, and cumulative deaths reported by CovidCounties against corresponding data reported by 8 different state Departments of Public Health. Data ranges vary by state; curated state data is available in the data file accompanying this manuscript.

An R^2^ of 1 was specifically found with respect to cumulative cases and deaths in Connecticut (Figure 2B), suggesting a shared common data source.

We compared the concordance of our data with that reported by Corona Data Scraper [https://coronadatascraper.com/], another widely used source of aggregated publicly-available COVID-19 timeseries data at the county level. We found very high concordance (R^2^ = 0.95-0.97) for deaths and cases respectively with no model bias (Figure 2C).

Lastly, we compared the concordance of our predicted hospitalizations, cases, and deaths from our dataset against data reported by 8 different State Departments of Public Health (Figure 2D). We noted some relative over-estimation of our estimated hospitalized counts against that actual data reported by multiple State Departments of Public Health. Nonetheless concordance was very high (R^2^ = 0.97-0.99).

Descriptive statistics on the New York Times data are provided in Table 1. 23 states reported cases with unknown counties of residence, however, in all states except Rhode Island these cases made up less than 4% of the total cases in that state (Table 1). The inability to map these cases to specific counties may explain some of the discrepancies between the New York Times data used in CovidCounties and the curated data from state public health departments and the Corona Data Scraper.

The data and tools incorporated into CovidCounties support the effectiveness of social distancing measures, consistent with several events that have occurred following the initial release of the website. South Dakota, one of six states which does not have a statewide shelter in place order (as of April 15, 2020), has experienced rapid case growth following an exposure at a meat plant (Figure 3A). This has accounted for more than half of the state’s cases^17^ as of April 15, 2020, with the fastest statewide doubling time of 4.5 days (Figure 3B). By contrast, states with early shelter in place times like Arizona on March 11, 2020, California on March 19, 2020, and Connecticut on March 20, 2020 (Figure 3A) have much slower doubling times of 19.3 days, 22.8 days, and 12.7 days respectively (Figure 3B).

**Figure 3:**
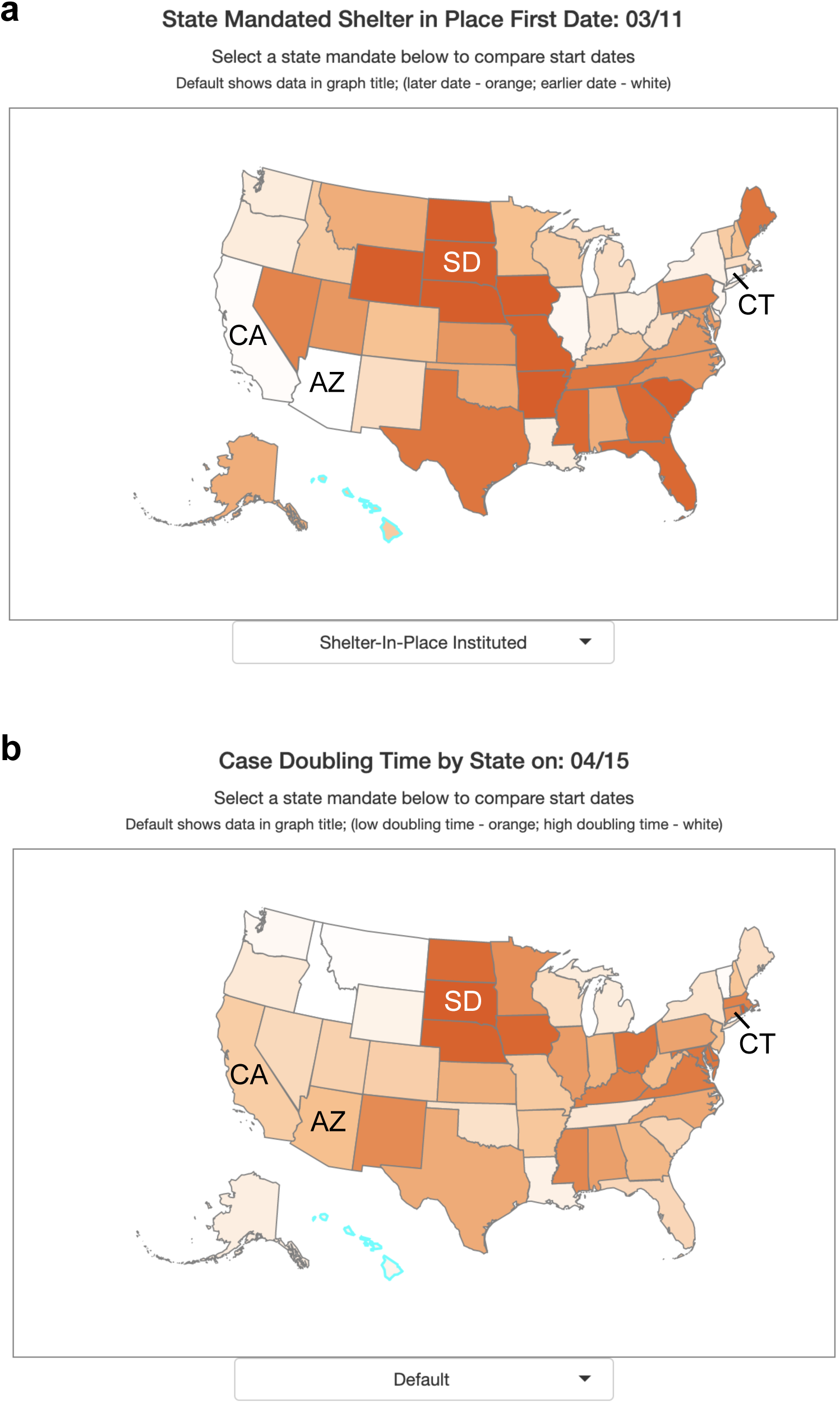
Effect of shelter in place orders on doubling time. A. States within the United States are color-coded by percentile of date to implement state mandated shelter in place. White indicates earlier dates (among states) while dark orange indicates later dates or no state mandate. B. States within the United States are color-coded by percentile of case doubling time on April 15, 2020. Dark orange indicates a fast doubling time (among states), white indicates a slow doubling time.

The web application located at covidcounties.org was first released to the public on April 3, 2020. It features two sections: a line plot depicting time-series trends in disease dynamics, and a map depicting geospatial relationships (Figure 4). The site has had over 15 thousand unique site in the first week as of April 11, 2020, most of whom accessed the website using a mobile device.

**Figure 4:**
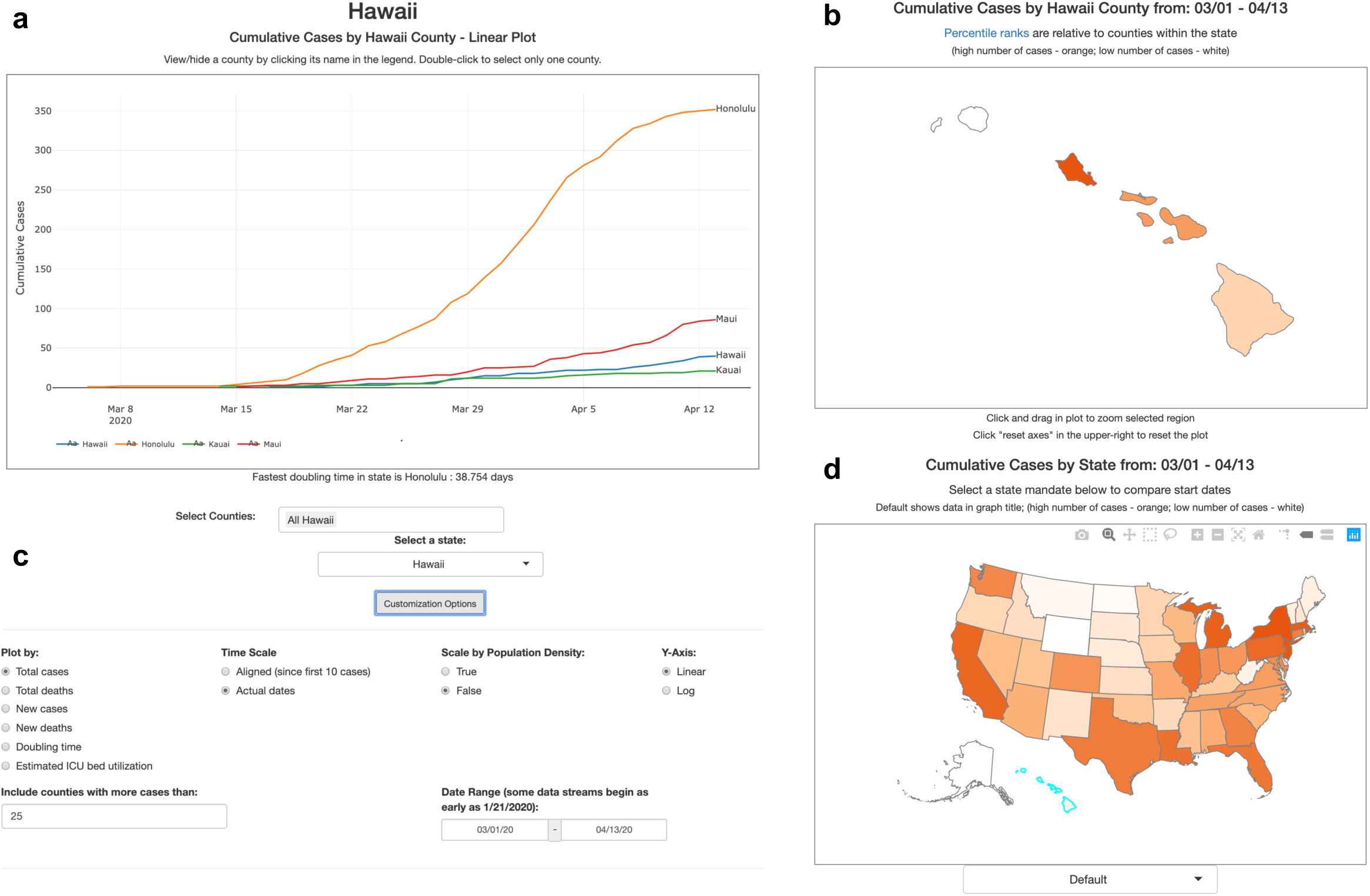
Overview of CovidCounties.org. A. The primary view of CovidCounties.org is the line plot view, depicting time-series trends by individual county. Depicted counties may be selected by single or double clicking the counties displayed in the legend. They may also be selected by typing in counties (including from outside of a given state) at the bottom. B. User-selected individual states are color coded according to the variable of interest (e.g. cumulative cases). Dark orange corresponds to the highest percentiles within the state, white indicates the lowest percentile. Hovering functionality displays statistics corresponding to a given county. C. Line plot views can be extensively customized, with features to enable axis recentering/scaling, count normalization, depiction of doubling time, and predicted ICU bed utilization. Individual state and United States plots update to reflect selected parameters where appropriate. D. States within the United States are color-coded by percentile according to the variable of interest (e.g. cumulative cases). Dark orange indicates relatively high percentile (among states), white indicates low percentile. Hovering functionality displays statistics corresponding to a given state. The dropdown menu below allows the user to change the view to depict timing that various social distancing policies were implemented: white indicates relatively early adoption (by percentile), dark orange indicates late or no current adoption.

## Discussion

The effective management of the COVID-19 pandemic has been hindered by both inaccurate data collection and reporting, as well as relative inaccessibility by non-data scientists. Taken together, these difficulties have impeded optimal policymaking by both government (imposing social distancing policies) and health systems (anticipating ICU utilization) alike. Consequently, responses across institutions have been highly variable and with varying degrees of success. To help address these gaps we developed covidcounties.org and performed the technical validation reported in this work.

The curation of COVID-19 case and death counts by The New York Times is an impressive effort by over 60 reporters to collect, curate and analyze a constantly growing and evolving dataset^3^. However, they acknowledge that the underlying data is extremely fragmented and comes from thousands of different sources at both the state and county levels and thus is inherently limited by accuracy, consistency, and timeliness. The New York Times notes that reported cases have been corrected mere hours after the initial report and there have been numerous instances where data has disappeared from databases without explanation. The New York Times has also chosen to count patients where they were treated rather than their place of residence and report on a number of geographic exceptions in their dataset (https://github.com/nytimes/covid-19-data) including the treatment of cities like New York City and Kansas City and the allocation of cases from cruise ships. Further, there are a subset of cases where the patient’s county of residence cannot or has not yet been identified which is generally a small fraction of a state’s total cases but can be a significant number in a small state like Rhode Island (Table 1).

Taken together, these subtleties of the data collection process imply that the COVID-19 data from The New York Times may not exactly agree with the numbers reported by various state and county Departments of Public Health. We quantified the consistencies between The New York Times COVID-19 data and county (Figure 2A, 2B) and state (Figure 2D) Department of Public Health data and found the datasets to be largely comparable. Based on the exact agreement, it seems likely that The New York Times is deriving their data for Connecticut directly from the Connecticut Department of Public Health (Figure 2B).

The comparison of our estimated hospitalized cases based on the simple model from Ferguson et al.^7^ with state (Figure 2C) and county (Figure 2B) reported hospitalizations revealed a systematic bias towards increased hospitalizations in our model. We suspect that this bias is due to a number of factors including time lags between the date of hospitalization and the results of testing, as well as miscalibration of the assumed 4.4% rate of hospitalization taken from the Ferguson model^7,8,11^.

With the advent of the COVID-19 pandemic we have observed a trend towards government agencies at the municipal, county, state, and national levels making their data increasingly accessible for re-use and therefore provide potential value. However, many of the most popular tools which are built upon this freely available data do not provide their source code for further development. The Johns Hopkins dashboard^2^, which receives more than 1.2 billion hits per day, has made their data publicly available^18^, however, the source code for their dashboard is not made available for further development by third parties. Similarly, the IHME dashboard^19^ which has been referenced by the White House for making policy decisions^20^ has had their dashboard peer reviewed^21^, however, their epidemiological model has yet to be peer reviewed^9^. While IHME provides open source code on their data aggregation process (https://github.com/beoutbreakprepared/nCoV2019) and some features of their model including the curve fitting of their projections (https://github.com/ihmeuw-msca/CurveFit), the whole dashboard is not open source. Additionally, many states and counties are using *Tableau*, a proprietary piece of software, to visualize COVID-19^22^ and as of 4/17/2020 there are 1,184 coronavirus dashboards on Tableau public^23^. While Tableau facilitates powerful data visualization, the software is not open source and requires a license for use. To promote further development of CovidCounties and fully leverage the available data we have implemented our website using the commonly used *R* and *Rshiny* frameworks, and made all of our source code freely available on github (https://github.com/vivical/ButteLabCOVID).

CovidCounties represents an improvement over existing dashboards in terms of both scope and granularity. Existing COVID-19 dashboards generally focus either on county level data within a particular state (primarily at a static timepoint) or at the state level across the United States. We have developed an intuitive tool that facilitates temporal comparisons between all counties in the US. However, we are inherently limited by the availability of data. While CovidCounties’ estimation of ICU needs at the county level allows for higher resolution allocation of resources compared to the widely used state level model from IHME (https://covid19.healthdata.org/united-states-of-america), zip code level data would further improve the value of these estimations for resource allocation. States like Maryland^24^, Arizona^25^, and South Carolina^26^ and counties like Johnson County, Kansas^27^, San Diego County, California^28^, and King County, Washington^29^ have already made zip code level data available. However, there are many states and counties that are hesitant to provide data of this granularity due to concerns over privacy thus highlighting the challenge of balancing privacy with public good.

A limitation of CovidCounties is the inherent dependence on publicly available data. To date, most states and counties are primarily providing case and death data with an increasing number also providing hospitalization data. However, there is a severe lack of testing information. The lack of testing data limits the ability to make inferences on the infection rate in the population and the improvement of model trajectories. It has also been proposed that there has been an under ascertainment of cases especially in the asymptomatic^30^, which can influence case rates. States and counties are continuously ramping up testing and this sudden availability of tests can artificially distort counts by attributing individuals who were infected previously to a later date due to an earlier shortage of tests. These numbers are further complicated by the wide variety of commercially available tests that rely on different technologies with varying sensitivity and specificity.

With its release, covidcounties.org represents a powerful open-source platform to empower non-data scientists to track the current trends of the COVID-19 pandemic at the county level to help facilitate policy and healthcare decisions which can help improve outcomes. We welcome volunteers (both technical and non-technical) to help us to further develop CovidCounties (https://covidcounties.org/buttelabcovid/www/volunteers.html).

## Usage Notes

A summary of the website features is available from the University of California, San Francisco [https://ucsf.app.box.com/v/Covid19Townhall041720]. A detailed tutorial illustrating use of the website is available on youtube.com (https://youtu.be/5OHDSpLv1kY).

## Code Availability

The website source code is available on github (https://github.com/vivical/ButteLabCOVID). A version-controlled *Docker* [ref] container is also available on dockerhub (https://hub.docker.com/r/pupster90/covid_tracker).

## Data Availability

Curated data on the state-wide implementation of social-distancing policies and curated validation data are hosted on datadryad.com (https://datadryad.org/stash/share/whGecW9DWYmoAVMDdAHNF0z712Vbxrj9YwI5QKRAWUs).

## Data Availability

Curated data on the state-wide implementation of social-distancing policies and curated validation data are hosted on datadryad.com

https://datadryad.org/stash/share/whGecW9DWYmoAVMDdAHNF0z712Vbxrj9YwI5QKRAWUs

## Contributions

PB and AJB jointly conceived the initial concept. DA, PB, ME, BO, and RV participated in data acquisition, software development and website deployment. AM, VAR, and TZ participated in manual collection and curation of the state governmental policy data. DA, AM, VAR, and TZ contributed to the initial draft of the manuscript. All authors performed editing for important intellectual content. PB, AJB, BO, and VAR performed project management and overall supervision.

## Acknowledgements

The authors kindly acknowledge Amazon Web Services for donating the necessary computing resources for website hosting.

